# Patterns of neurodegeneration in dementia reflect a global functional state space

**DOI:** 10.1101/2020.11.09.20228676

**Authors:** D. Jones, V. Lowe, J. Graff-Radford, H. Botha, D. Wiepert, M.C. Murphy, M. Murray, M. Senjem, J. Gunter, H. Wiste, B. Boeve, D. Knopman, R. Petersen, C. Jack

**Author notes:** Correspondence to: David T. Jones, Mayo Clinic, 200 First Street S.W., Rochester, MN 55905.

## Abstract

Disruption of mental functions in Alzheimer’s disease (AD) and related disorders is accompanied by selective degeneration of brain regions for unknown reasons. These regions comprise large-scale ensembles of cells organized into networks required for mental functioning. A mechanistic framework does not exist to explain the relationship between clinical symptoms of dementia, patterns of neurodegeneration, and the functional connectome. The association between dementia symptoms and degenerative brain anatomy encodes a mapping between mental functions and neuroanatomy. We isolated this mapping through unsupervised decoding of neurodegeneration in humans. This reflected a simple information processing-based functional description of macroscale brain anatomy, the global functional state space (GFSS). We then linked the GFSS to AD physiology, functional networks, and mental abilities. We extended the GFSS framework to normal aging and seven degenerative diseases of mental functions.

**One Sentence Summary:** A global information processing framework for mental functions links neuroanatomy, cognitive neuroscience and clinical neurology.

## Main Text

Mapping biological functions to their anatomic substrates has been a central theme throughout medicine. At the core of the clinical practice of neurology is the localization of a particular clinical deficit to an anatomic substrate in the nervous system. Localizing limb strength to a lesion in the nervous system is usually straightforward, but in dementing neurodegenerative disorders of the brain, clinical symptoms manifest as selective impairments in mental functions. Cognitive psychology describes these mental abilities using terms such as perception, emotion, memory, social cognition, language, and executive function. Localization of these functions is coarse grained and poorly understood as there is no framework describing the high-level relationships between anatomy, brain dynamics, and mental functioning to guide the clinical approach to these common conditions. Lack of understanding of this biology also precludes the development of precise neurodegenerative disease models that include physiology related to dynamic large-sale functional brain systems like the default mode network (DMN) (1-3). In order to bridge this divide, a mapping between concepts in computational neuroscience, clinical neuropsychology, and neurology is required. Such a framework will provide the foundations for improved clinical management of disorders of brain function.

From a computational neuroscience perspective, the diverse cognitive functions selectively degraded by Alzheimer’s disease (AD) and related disorders, arise from global integration of the ongoing microscale and mesoscale dynamic functional operations occurring within a relatively fixed spatial anatomy. In this respect, high-level mental abilities emerge from the dynamic global integration of local integrators (4). These globally integrated units can be modeled as large-scale network topologies embedded in hierarchical adaptive network architecture (5). Emergent properties of particular network topologies could then facilitate specific classes of mental abilities. Therefore, a complete spatial mapping of these macroscale network configurations would represent a functional-anatomic mapping of the biology involved in dynamically optimizing perception, cognition, and behavior (6, 7) that are selectively targeted in neurodegenerative diseases of the brain (8). Indexing such a large number of potential network configurations seems like an intractable problem on the surface, but this repertoire of network topologies has recently been described by trajectories on a low-dimensional manifold, and apparent neurotransmitter-modifiable flow through this manifold supports diverse mental abilities (7). We refer to this low-dimensional manifold as a global functional state space (GFSS). Theoretically, each point in this state space would represent a particular global network topology that optimally supports a specific mental ability, while impairment in a particular class of high-level mental functions would correspond to altered dynamics involving a portion of state space associated with that function. Clinically, such limited flow across a portion of the GFSS would be characteristic of a particular dementia syndrome. Pathologically altered large-scale dynamics would explain the observed similarity between degenerating brain anatomy and functional connectivity patterns, as well as the selective clinical impairment of a particular high-level mental function. In the current study we find evidence for these predicted relationships, and incorporate them into a mechanistic model explaining the relationships between neurodegenerative patterns, functional connectivity, and clinical symptoms. This is accomplished within a framework that emphasizes functional modes of degeneration within a continuous dynamic functional state space rather than molecular spreading along functional connections.

The neurobiology that allows for AD and related disorders to selectively target particular mental abilities or large-scale brain networks, while sparing others, is unknown. There are prominent individual differences in this selectivity leading to variable cognitive symptoms among types of AD dementia, such as the typical late life amnestic dementia syndrome and the younger-onset visual or dysexecutive variants (9), resulting in a “paradox of syndromic diversity” (10). Characterizing the factors related to this paradox, and individual variability in general, will inform our understanding of the underlying neurobiology driving syndromic variability and mental functions (11). Recent investigations of individuals with typical AD and those with the visual variant of AD showed that they were indistinguishable at the molecular level (12, 13), but they can be distinguished at the network level (14, 15) or by large-scale tau patterns that resemble the spatial anatomy of functional brain networks (16). This suggests that inter-individual variability in mental abilities affected by AD is partly driven by inter-individual differences in the macroscale functional pathophysiology of AD, as opposed to purely at the molecular level. We have recently proposed a theory of AD pathogenesis that implicates large-scale network dynamics in disease pathogenesis alongside microscale misfolding of proteins (3, 16, 17) that is in-line with general theories of network dysfunction in neuropsychiatric disease (18). In order to test and improve upon AD models that include large-scale network physiology, a more complete model of the physiology of mental abilities and their relationship to brain networks and neuroanatomy is essential. This requires a framework that spans computational neuroscience, clinical neuropsychology, and neurology. Theoretically, the GFSS can provide such a framework, but evidence for selective degeneration of modes/regions of the GFSS does not yet exist.

In our proposed GFSS framework, the selective functional impairment seen in an individual with AD results from impaired dynamics in a particular portion of the state space manifold, or degenerative dynamics within a functional mode of operation (3). In other words, the specific pattern of global dysfunction in an individual would represent a particular parameterization of disease pathophysiology at this global scale. In this disease model, individuals with neurodegenerative diseases represent unique “lesion studies” disrupting dynamics of GFSS functional modes. We looked for evidence of this to support a GFSS framework of neurodegenerative diseases of brain function. We hypothesized that the inter-individual differences in neurodegeneration across the AD spectrum could be decoded to yield the GFSS, providing a direct link between AD pathophysiology and the low dimensional manifold of large scale brain organization. We further hypothesized that this GFSS would be related to functional imaging literature and be applicable to interpreting clinical data across normal aging and dementia syndromes.

There is no existing method of determining how impaired flow through the GFSS could be encoded by measures of brain function in AD patients. However, if flow through the GFSS is fundamental to the macroscale neurobiology of AD, then it should be possible to use a form of latent variable analysis to decode functional imaging data from patients, construct a representation of the GFSS, and subsequently determine the extent to which AD effects are predicted by the GFSS (Fig. S1). In this study, we construct an estimate of the GFSS and examine its relationship to fundamental features of AD. This is accomplished through four main investigations: 1) human data (N = 423) is used to derive the GFSS via an unsupervised pattern analysis and latent space representation of global glucose uptake across the AD clinical spectrum, 2) mental functions are mapped to the observed GFSS using a functional meta-analysis and compared to similar mappings obtained using functional connectivity data, 3) application and validation of the predictive ability of the observed GFSS in a large multi-site study (N = 410), and 4) additional clinical validation of the functional-anatomic mapping by projecting data from normal aging (N = 1,121) and clinically defined dementia syndromes (N = 291) selectively targeting memory, executive functions, language, behavior, movement, perception, semantic knowledge and visuospatial abilities. Our results provide further validation a GFSS framework and demonstrate its translational potential to improve the practice of neurology through advances made in neuroimaging and computational neuroscience.

### Patient Selection

In order to ensure that the GFSS was disrupted in the individuals included in our investigation, we selected patients with evidence of cognitive impairment, defined here as a clinical dementia rating scale greater than zero. In this patient population, we aimed to investigate brain physiology that would be sensitive to changes in the GFSS that can be reliably measured on the single-subject level. Therefore, we studied glucose uptake measured by F18-fluorodeoxyglucose (FDG) positron emission tomography (PET), a widely used functional imaging modality in routine use in our clinical practice currently. In the current research framework for AD, FDG-PET is considered a biomarker of neurodegeneration (*19*). We limited our FDG-PET analysis to individuals who had evidence of microscale AD pathophysiology (i.e., elevated beta-amyloid PET). While this focuses our investigation to individuals with a microscale element of AD pathophysiology by definition (*19*), it does not preclude other co-morbid conditions and therefore allows for a sampling of the complete spectrum of beta-amyloid associated cognitive impairment. We identified 423 patients that meet these inclusion criteria (Table 1). The characteristics of the validation cohort form the Alzheimer’s Disease Neuroimaging Initiative (ADNI) are also displayed in Table 1.

**Table 1.** Mayo and Alzheimer’s Disease Neuroimaging Initiative (ADNI) Sample Characteristics APOE e4+ – carriage of an APOE-ε4 allele; CDR– Clinical Dementia Rating Scale; CDR-SOB – Clinical Dementia Rating Scale Sum of Boxes; MMSE – Mini-Mental State Examination

|  | Mayo | ADNI | P-value |
| --- | --- | --- | --- |
| N | 423 | 410 |  |
| Age (median [Q1,Q3]) | 77.4 [69.1, 83.5] | 74.5 [69.6, 79.4] | 0.001 |
| Male (%) | 244 (57.7) | 223 (54.4) | 0.375 |
| Education (median [Q1,Q3]) | 15 [12, 17] | 16 [14, 18] | <0.001 |
| APOE $\epsilon 4+$ (%) | 243 (61.7) | 283 (69.2) | 0.030 |
| CDR (%) |  |  | <0.001 |
| 0.5 | 274 (64.8) | 310 (75.6) |  |
| 1 | 106 (25.1) | 96 (23.4) |  |
| 2 | 39 ( 9.2) | 4 ( 1.0) |  |
| 3 | 4 ( 0.9) | 0 ( 0.0) |  |
| CDR-SOB (median [Q1,Q3]) | 3.0 [1.0, 5.5] | 2.0 [1.0, 4.4] | 0.004 |
| MMSE (median [Q1,Q3]) | 24 [21, 27] | 26 [24, 28] | <0.001 |
| FDG (median [Q1,Q3]) | 1.25 [1.09, 1.40] | 1.16 [1.05, 1.28] | <0.001 |

### Latent Space Derivation of the GFSS

Individual variability in large-scale patterns of glucose uptake in these patients can be conceptualized as a parameterization of amyloid associated AD neurobiology at a global scale (*11*). We decoded this parametrized AD pathophysiology by performing a form of multivariate pattern analysis on a between-subject level that we refer to as Between-subject variability Projection and Reduction (BPR) to emphasize the importance of sample selection and patient factors for decoding pathophysiology.

The BPR algorithm leverages widely used analytic techniques to identify global patterns of between subject covariance. As we applied it to our imaging data, it is a higher-dimensional computational equivalent of the 2-D eigenfaces facial recognition algorithm as implemented by Turk and Pentland (*20*). Unsupervised linear (singular value decomposition) and non-linear (Laplacian eigenmaps) methods for the manifold decoding step performed similarly in our data, but a full sampling of the parameterization of the GFSS, via between subject variances, is required in order to replicate the decoding. This is because BPR, and related analyses, of FDG-PET images from a disease class will index meaningful global features of altered glucose uptake caused by the pathophysiologic process of interest in the patient population being studied. In the population we studied, this algorithm produced a low dimensional linear basis-set of eigenbrains or EBs (Fig. 1), that describes 95% of the variability in the FDG images (Figs. S2-S5). These EBs describe global patterns of variation in glucose uptake among the group that index highly meaningful functional brain properties relevant to AD biology. Further analyses outlined below supports our hypothesis that these patterns describe information processing aspects of the GFSS of interest in this study.

**Fig. 1.**
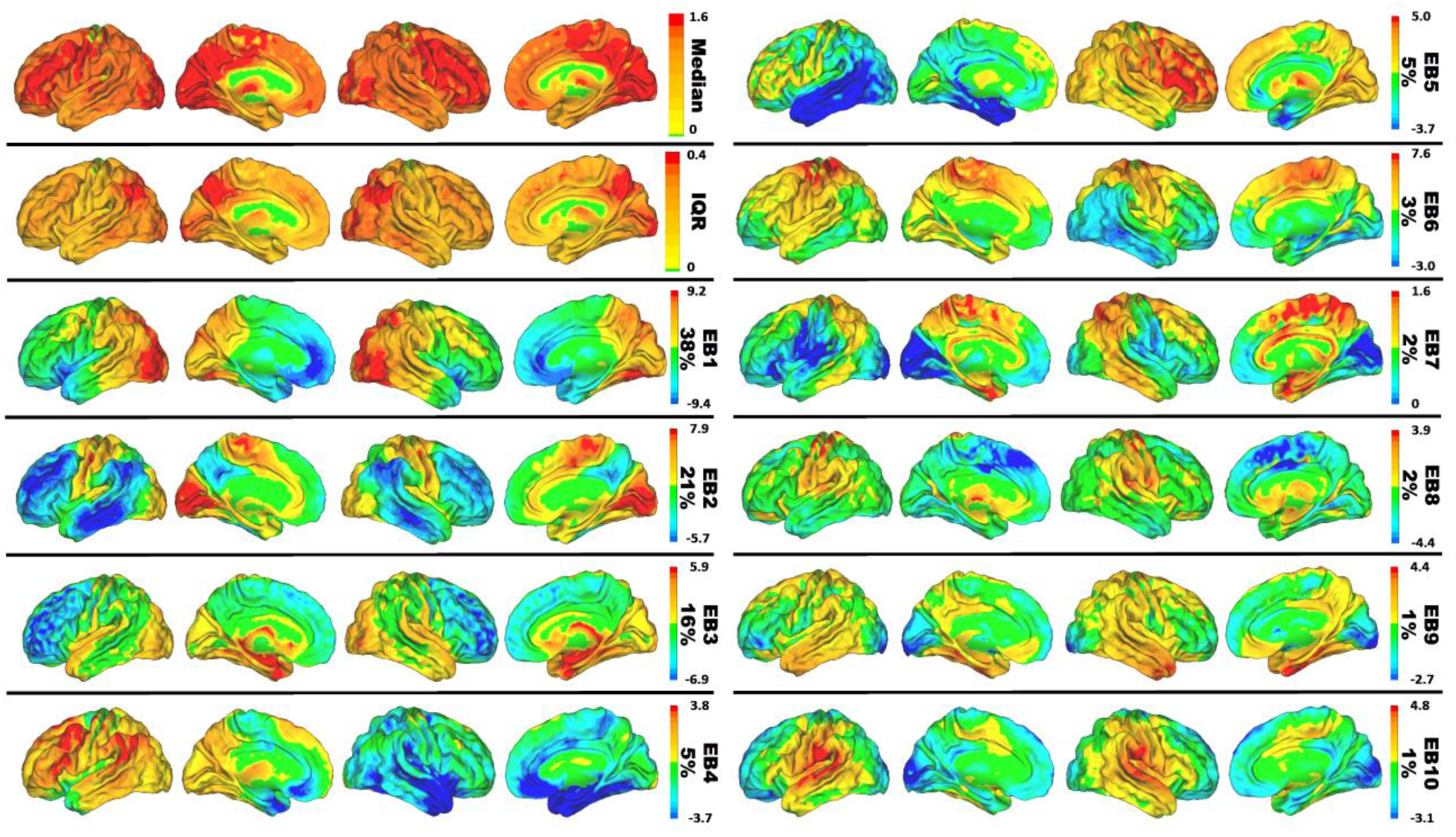
Eigenbrain decomposition of glucose uptake in 423 Alzheimer’s patients reveals a low-dimensional set of large-scale patterns that explain 95% of the variance among patients. Surface renderings of median, interquartile range, and eigenbrain intensities for the first 10 eigenbrains are displayed. The percentage of variance explained by each is listed to the right of the color bar.

### Functional Mapping of the Anatomy Described by the Glucose Eigenbrains

We used a NeuroSynth (www.neurosynth.org) (*21*) functional topic terms (*22*) based decoding (*23*) as a common framework to compare the functional anatomy captured in this study to the existing functional MRI literature in a similar manner as Shine, et al. 2019 (*7*) and Margulies, et. al. 2016 (*24*), allowing for a common understanding of these diverse findings in the same meta-analytic functional terminology. The functional topic term decoding for a single topic across all 10 EBs can also be used as an embedding of that topic in the GFSS coordinate system. The coordinates of that GFSS embedding can then be used as EB weights in a linear anatomical reconstruction of that functional topic (Fig. 2). The linear combination of the smooth gradients described by the EBs produce global patterns associated with each functional topic. Peak values in these reconstructed maps correspond to regions of peak activation associated with brain patterns observed during performance of these tasks. These topic term embeddings can also be used as ‘functional waypoints’ to aid in interpreting functional correlates of large-scale anatomic patterns of disruption in patients. In order to do this on a single subject level across functional topic terms, we linked values in the GFSS coordinate system continuously to the functional imaging literature via a full topic term decoding of each EB (Fig. 3) and then embedded individual subjects into this well characterized functional-anatomic coordinate-based framework (Fig. 4).

**Fig. 2.**
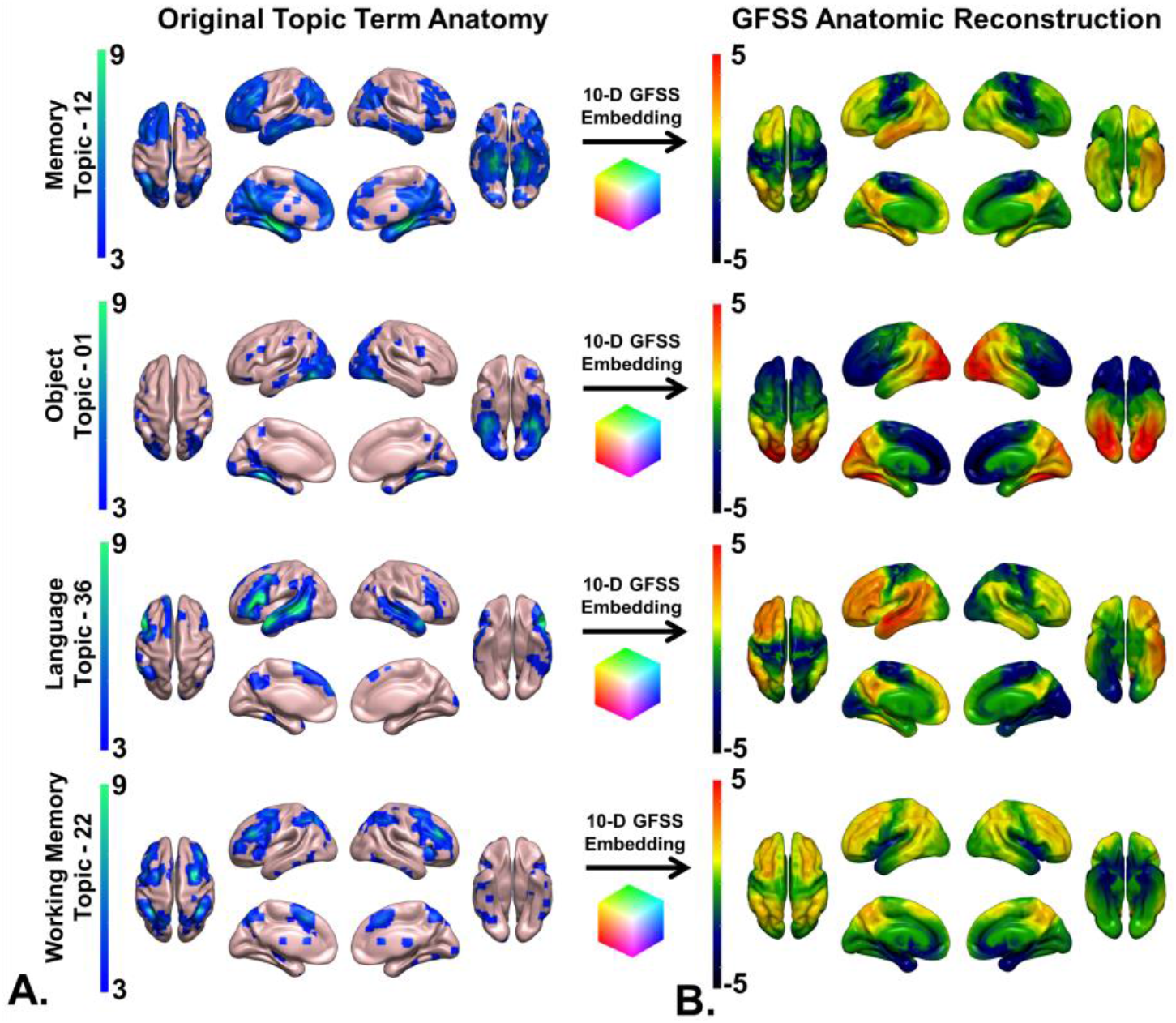
Embedding large-scale brain patterns associated with mental function topics. A) NeuroSynth (http://neurosynth.org/analyses/topics/) anatomic maps for four topics related to AD clinical syndromes are displayed on surface renderings. B) The GFSS embeddings of these topics were used to generate the anatomy associated with that point in the GFSS manifold and projected onto surface renderings. This demonstrates faithful representation of these anatomic patterns associated with mental topics in the GFSS.

**Fig. 3.**
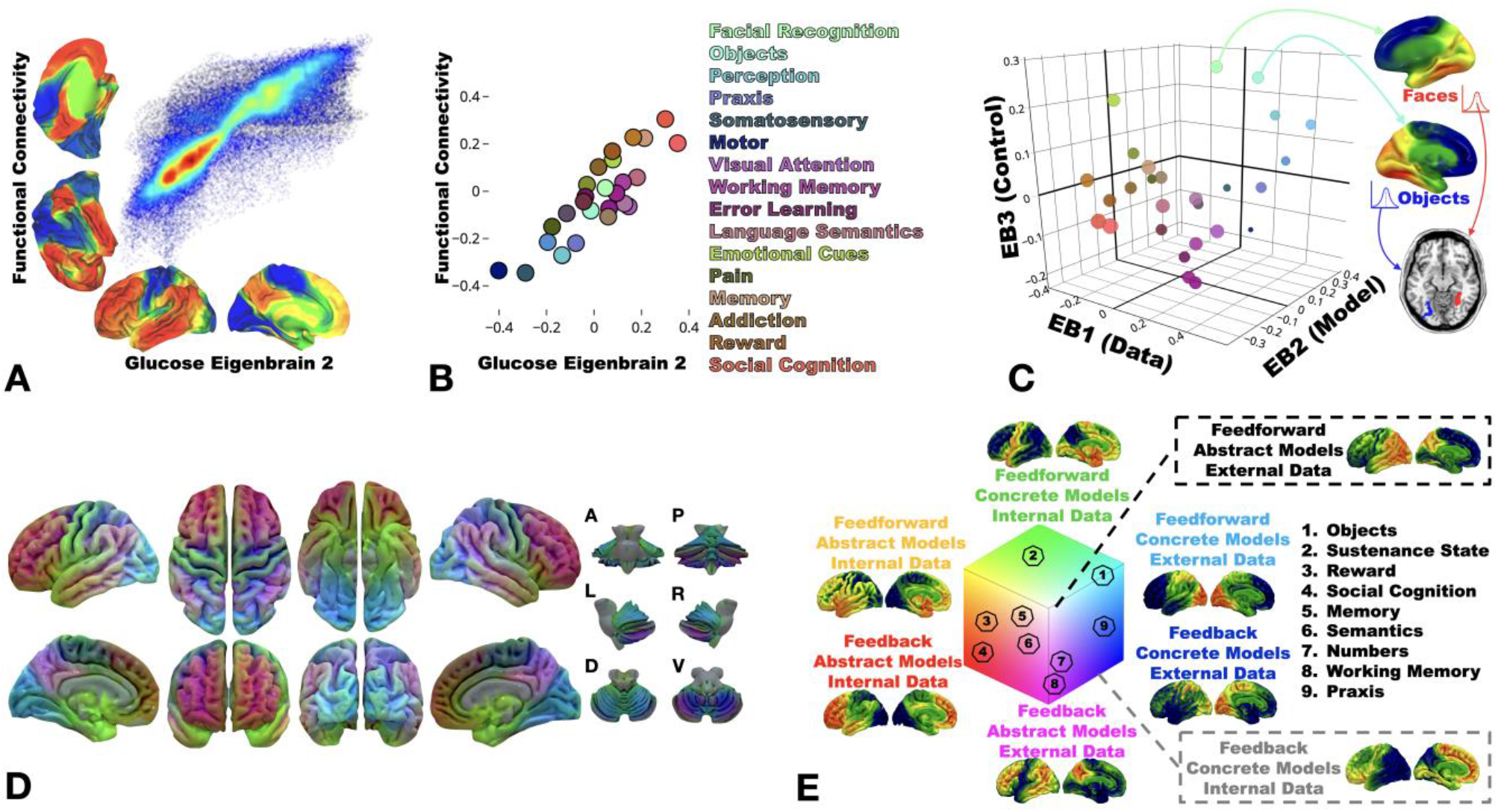
Information processing and the global functional state space. A) Joint histogram between the principle axes of functional connectivity (13) and glucose EB2. B) NeuroSynth decoding of the principle axes of functional connectivity versus glucose EB2 decoding. Select topic terms are color-coded on the right (color coding is the same as in C). C) Scatter plot of topic term decoding for glucose EB1-3. For the color-coding, each EB decoding was used as a RGB channel (EB1 = Blue, inverted polarity EB2 = Red, EB3 = Green). Radius of the points encodes depth along EB2. Generated anatomy using the 10-D EB decoded coordinates (Table S3) for faces and objects are displayed on the right and left hemisphere (respectively) surface renderings. Below these, axial brain slice with the peak voxels from the faces and objects anatomic projection overlaid highlighting the fusiform face area (red) and visual word form area (blue) respectively encoded at these points. D) The same RGB color-coding was applied in a voxel-wise manner using the intensities from EB1-3 producing a continuous functional parcellation of brain anatomy along these gradients. E) The state space representation of the same color mapping with the approximate location of nine cognitive topic terms from panel C overlaid and numbered. A surface rendering of the anatomic correlates, generated from linear combinations of EB1-3 weighted by the position in state space, for the eight extremes of the cube are displayed near the portion of state space represented. A-anterior; P-posterior; L-left lateral; R-right lateral; D-dorsal; V-ventral

**Fig. 4.**
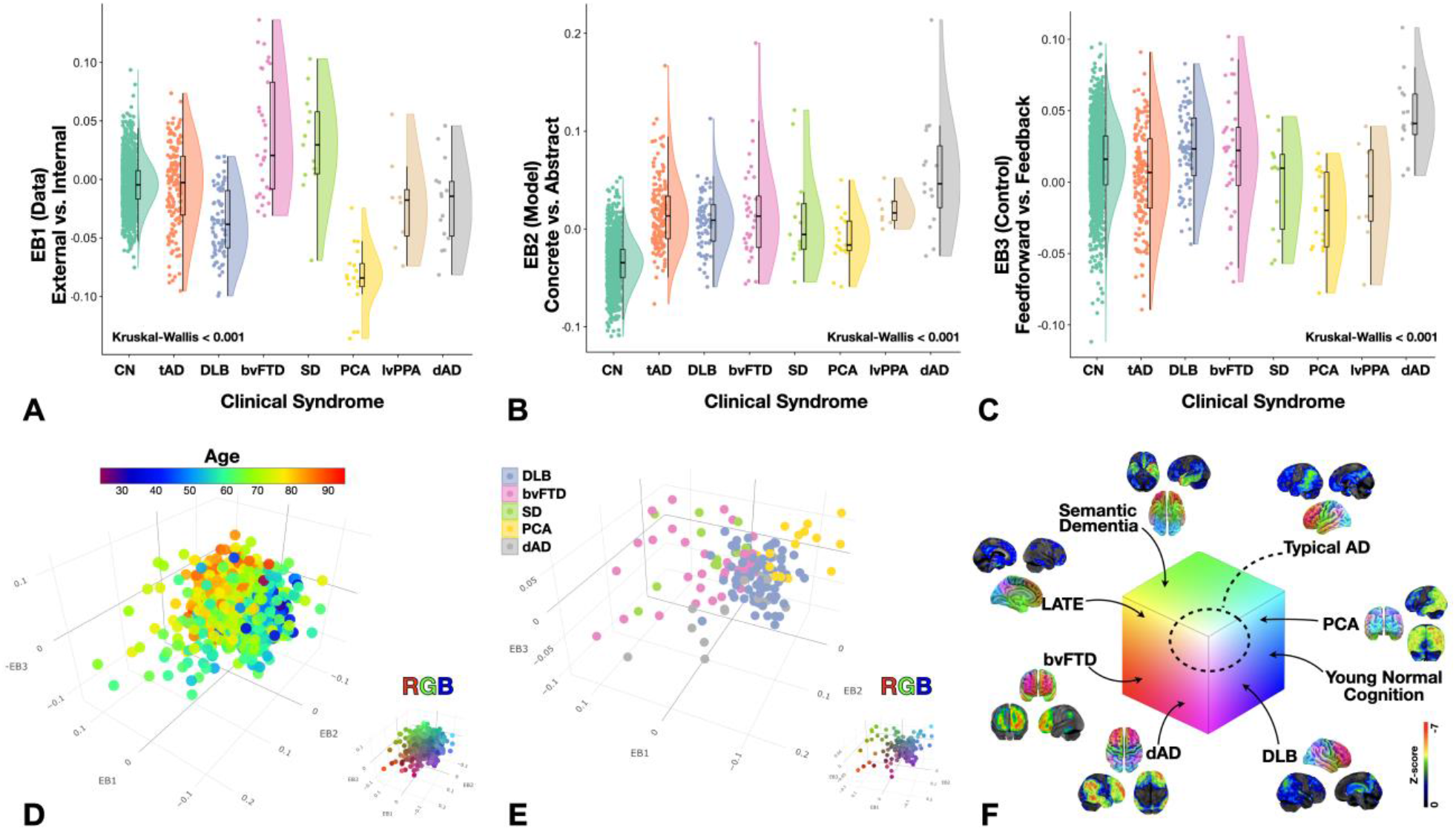
GFSS across normal aging and seven clinical dementia syndromes. Rain cloud plots (28) with data distribution and jittered raw data points for each subject on either side of boxplots of eigenvalues for A) EB1, B) EB2, and C) EB3 for cognitively normal amyloid negative individuals across the aging spectrum (CN), typical AD (tAD), dementia with Lewy bodies (DLB), behavioral variant of frontotemporal dementia (bvFTD), sematic dementia (SD), posterior cortical atrophy (PCA), logopenic variant of primary progressive aphasia (lvPPA), and dysexecutive AD (dAD). D) Scatter plot for the first 3 EBs for all of these subjects with age color mapping showing the youngest individuals (blue) at the opposite extreme from the oldest individuals (red). The same plot with RGB color mapping for reference to other figures is inset in the bottom right. E) Scatter plot for the first 3 EBs for 5 dementia syndromes highlighting the differential mapping across the GFSS with clinical syndromes coinciding with predictions made by the functional mapping in Fig. 3. The same plot with RGB color mapping for reference to other figures is inset in the bottom right. F) The same RGB GFSS color mapping used in Fig. 3 indicating the anatomy and GFSS location for each clinical group (including an example of limbic-predominant age-related TDP-43 encephalopathy [LATE] (29)). A representative single subject clinical FDG-PET (Cortex ID, GE Healthcare software) with z-scores relative to age matched controls color-coding the degree of hypometabolism for one patient from each group is also displayed.

In this cohort, the first three EBs account for 75% of the variance and are related to hemispherically symmetric orthogonal axes of brain function that capture the majority of the hypothetical GFSS. Therefore, we focused on presenting the results for decoding and characterizing these three EBs. The functional axis captured in EB2 (Fig. 3A) was nearly identical to the principle gradient defined by Margulies *et. al*. (2016) using functional connectivity data from cognitively unimpaired individuals (*24*). The meta-analytic functional topic terms based decoding for EB2 and the same decoding of the principle gradient were highly correlated (Fig. 3B). This EB fully indexes the glucose uptake in the principle gradient of macroscale cortical organization, characterized at one extreme by heteromodal association cortex (centered on DMN regions) and on the other extreme by primary sensory and motor regions. This fundamental organizing feature of brain function was first observed in FDG-PET (*1*), subsequently identified in patterns of functional connectivity (*25*), and also shown to be impaired in AD (*2*). Features of this pattern (e.g., sparring of the sensorimotor strip) are also routinely used by clinicians when interpreting FDG scans from patients (*26*). The fact that global variation in glucose metabolism in AD takes place along this and other macroscale functional gradients supports our hypothesis that AD alters flow through a low dimensional, functional manifold which captures large-scale network configurations underlying mental functions (*7*).

This structural-functional mapping of the EBs can be compactly represented and visualized in a three-dimensional approximation of the GFSS using the first three eigenbrains. This can be done using a latent space coordinate system (Fig. 3C and Fig. 3E), or in anatomic space (Fig. 3D). The RGB color map of the anatomic representation clearly demarcates functionally meaningful brain parcels based on the patterns of continuous variation in the gradients of the first three eigenbrains. This produces analogous results to defining brain parcels based on regional variation in cytoarchitectonics (*27*).

Each of the GFSS axes, or latent variables, can be conceptually simplified and dichotomized via axis polarity informed by this brain-behavior mapping (EB1: data source [internal vs. external], EB2: model form [abstract vs. concrete], and EB3: control type [feedback vs. feedforward]). These conceptual labels are provisional based on the relations between functional topic term mappings, but they are in line with the anatomic connectivity, functional activation, and clinical symptoms described here. These labels also compliment current theories describing the functions of the nervous system in computational terms and provide a conceptual link between computational neuroscience and clinical neurology.

The three-dimensional approximation is hemispherically symmetrical, but EB4 and EB5 can be included to capture breaks in symmetry and cumulatively explain 85% of the variance in the dataset. Naturally, the relative variance explained depends on the phenotypic composition of the cohort studied, in line with the BPR formulation. For example, EB5 captures hemispheric asymmetries in the left temporal lobe, including regions relevant for language functions, and eigenvalues were higher for the patients diagnosed with the language-variant of AD relative to the rest of the cohort, two-sample t(421) = 3.69, p < 0.001. The topic terms based decoding of all 10 EBs and the principle gradient from Marguiles et. al. (2016) (*24*) are presented in Table S1.

### Predictive Modeling of Factors Related to AD

Together, the set of 10 EBs could be used to predict key demographic, imaging, clinical, and pathologic variables associated with the effects of AD (Table 2). In other words, indexing variation in glucose uptake in global brain systems relevant for mental functions is highly predictive of key effects of AD biology on an individual.

**Table 2.**
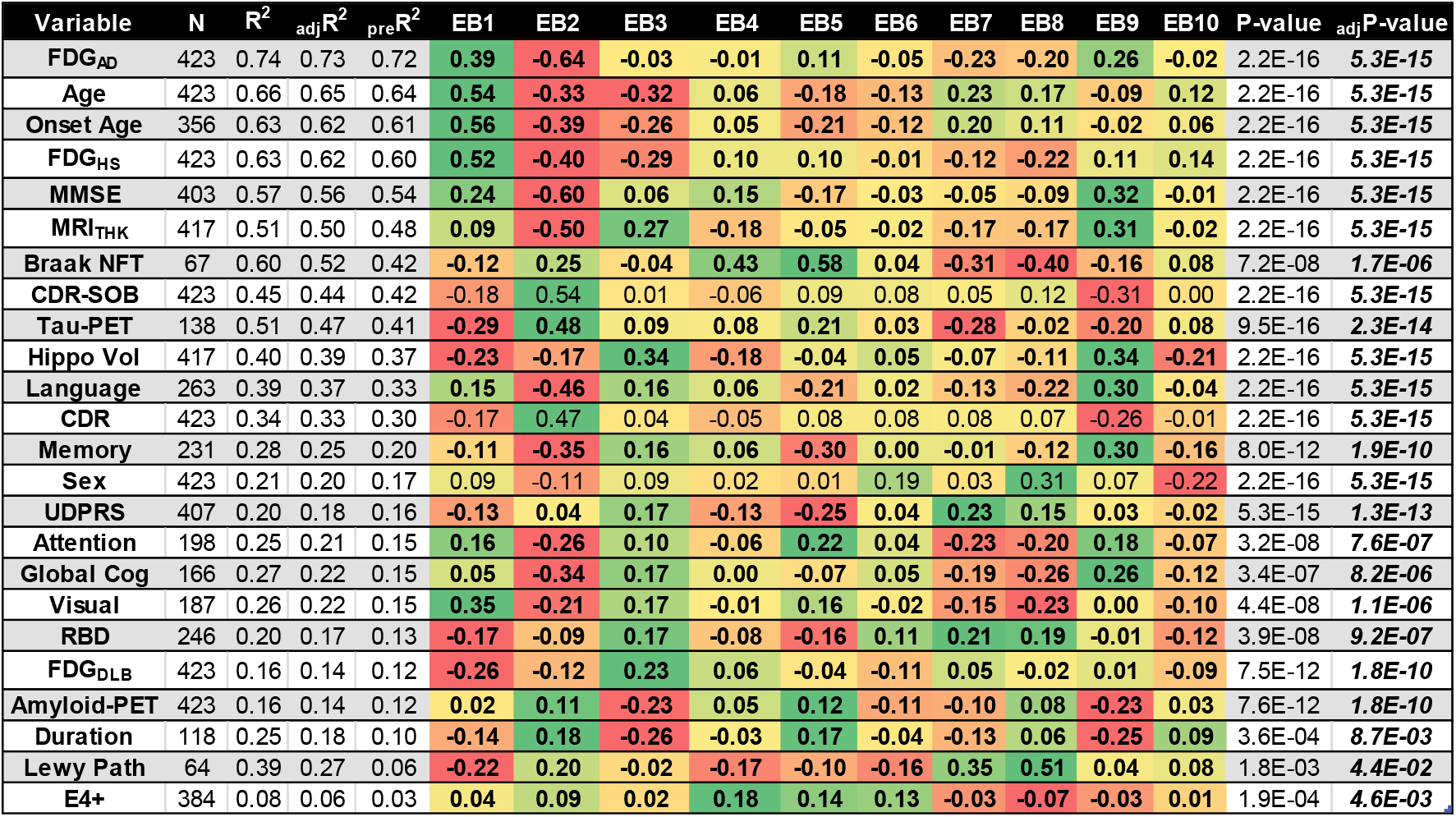
Predictive Models in the Mayo Cohort for Key Effects of Alzheimer’s Disease. For each of the dependent variables, the first 10 eigenvalues were used as predictors in a multivariate linear model. The R^2^, adjusted R^2^, predicted R^2^, color-coded magnitude and sign of standardized beta coefficients, p-value, and Bonferroni adjusted p-values are displayed for each model. FDG_AD_ – FDG SUVR in AD signature regions; FDG_HS_ – FDG SUVR in hippocampal sclerosis signature regions; FDG_DLB_ – FDG SUVR in DLB signature regions; MMSE – Mini-Mental State Examination; MRI_THK_ – MRI thickness in AD signature regions; Braak NFT – Braak neurofibrillary tangle stage; CDR– Clinical Dementia Rating Scale; CDR-SOB – Clinical Dementia Rating Scale Sum of Boxes; Hippo Vol – Hippocampal volume; UPDRS - Unified Parkinson’s Disease Rating Scale; RBD – REM sleep behavior disorder; Duration – disease duration; E4+ - carriage of an APOE-ε4 allele

### External Validation of Predictive Modeling

We next validate the predictive ability of mapping dysfunction in the GFSS in an independent cohort. Using the associations in this basis set to predict the age of patients from an independent database (N = 410) available as part of the Alzheimer’s Disease Neuroimaging Initiative (Table 1), we achieved a mean absolute error of 5.1 years using a linear 10 EB model. Similar results were obtained predicting other variables in the dataset related to glucose uptake, cognition, and disease severity, with peak prediction performance achieved with models using 8-20 EBs (Fig. S8 and S9). This strong predictive ability is suggestive of a key role for the GFSS in the expression of AD pathophysiology within an individual and serves as a robust validation of our results in a multisite study.

### Clinical Validation of the GFSS Framework

We embedded a large cohort of Mayo Clinic participants in the GFSS framework using the eigenbrains derived from only the 423 individuals with amyloid associated cognitive impairment (Fig. 4). See the Materials and Methods for exploration of the effect of cohort on eigenbrain definition (Fig. S5 and S6).

This cohort included cognitively unimpaired individuals with negative amyloid-PET scans (n = 1,121) across the age spectrum (median age [q1, q3] = 65 [57, 74], range = 30-93) and seven clinically defined age-associated dementia syndromes: typical Alzheimer’s disease (tAD, n = 137), Dementia with Lewy Bodies (DLB, n = 72), behavioral variant of frontotemporal dementia (bvFTD, n = 33), semantic dementia (SD, n = 11), posterior cortical atrophy (PCA, n = 15), logogenic variant of primary progressive aphasia (lvPPA, n = 8), and dysexecutive Alzheimer’s disease (dAD, n = 15).

Each clinical syndrome could be uniquely characterized by their distribution along the first three coordinates of the GFSS in a manner reflecting their distinguishing clinical features (Fig. 4A-C). All of the cognitive dementia syndromes differed from cognitive aging in terms of brain regions involved in abstract model formation (Fig. 4B). Both PCA and DLB displayed characteristic abnormalities in brain regions abstractly modeling information form external data sources (Fig. 4A), but brain regions important for feedforward control were more abnormal in PCA relative to DLB (Fig. 4C). Subjects with bvFTD and SD displayed characteristic abnormalities in brain regions abstractly modeling internal data sources (Fig. 4A), but SD involved more feedforward control brain regions relative to bvFTD (Fig. 4C). Both lvPPA and dAD groups showed the most extreme abnormalities in abstract modeling brain regions relative to other dementia groups, but dAD subjects were characteristically more impaired in brain regions supporting feedback control, in line with their characteristic working memory impairment (*9, 16*). Typical AD is characterized by being in the middle of these extremes.

## Discussion

Neurodegeneration in dementia syndromes can be conceptualized as occurring along a continuous low-dimensional manifold that spans the macroscale functional-anatomic organization of the brain, or global functional state space (GFSS). The coordinates in this state space encode macroscale anatomy that supports the cognitive functions which are selectively impaired by neurodegenerative diseases of the brain (Figs. 2-4). This macroscopic description of high-level functional organization compliments smaller scale anatomical mappings of lower-level brain functions (e.g., orientation columns in primary visual cortex) and highlights the multiscale nature of the brain’s functional architecture and its dysfunction in neurologic illness. The high predictive ability of the GFSS embedding for all major readouts of AD effects on an individual (Table 2), suggests that the global functional physiology of the brain is a key parameter influencing the expression of AD within an individual. These latent factors, related to information processing (Fig. 3), were decoded from patterns of glucose uptake in patients with AD, but are readily able to represent functional activation patterns from cognitively normal individuals. They are also able to capture clinically relevant patterns of variability across seven dementia phenotypes (Fig. 4). This same manifold can be found from decoding metabolic patterns across the aging and dementia spectrum (Fig. S6). Together, these facts lend support to complex systems based pathophysiologic models for all neurodegenerative diseases that integrate macroscopic functional physiology with microscopic cellular and molecular physiology (*3, 16, 17*).

Selective vulnerability of certain brain anatomic structures, large scale-brain networks, and the cognitive functions these networks and anatomy support, is a hallmark of all neurodegenerative diseases. This leads to a characteristic mapping between clinical phenotype, structural anatomy, and brain networks (*8*). The GFSS interpretation recasts these relationships in terms of degeneration in modes of brain functioning along a continuous state space. A mechanistic model of this selective degeneration requires an understanding of the physiology that allows static brain structure to support dynamic reconfiguring of functional operations in response to current high-level functional demands through coordination of spiking activity in large populations of neurons (*4*). In other words, a framework bridging cognitive computational neuroscience and clinical neurology is needed. The GFSS framework represents an important step in that direction conceptually, in addition to demonstrating immediate clinical utility (e.g., differentiating the most common causes of neurodegeneration form normal aging).

The GFSS is able to capture information represented discretely by meta-analysis of functional MRI (Fig. 3) or discrete patterns of degeneration in patients with dementia (Fig. 4), but in a continuous manner, suggesting a wide applicability of this latent space beyond FDG images in individuals with AD. Degenerative diseases are an important population to study neurodynamics because the pathophysiology in these conditions must selectively impair modes of function within the GFSS when they limit particular high-level functional abilities (memory, social cognition, executive control, semantic knowledge, visuospatial processing, etc.). Therefore, they represent “lesion studies” of functional neurodynamics. We did find it striking that our study of only AD associated cognitive impairment revealed such a comprehensive state space robust to the sample characteristic and methods used to derive it (Figs. S4-S7). This phenomenon likely relates to the wide clinical phenotypic variability in AD, the low dimensional nature of the GFSS, and the necessary dependencies within the neurodynamics regulating flow through this state space.

It is remarkable that three brain patterns related to high-level informational processing (information source [EB-1], model type [EB-2], and control mode [EB-3]) are sufficient to explain the majority of the variability in degenerative pattern formation in AD and related disorders. Necessarily, these eigenbrains also encode patterns observed in the functional MRI literature. We believe this occurs because the macroscopic functional properties encoded by the GFSS index state variables of the brain’s complex adaptive information processing system at a scale relevant for high-level cognitive functions. These cognitive functions are routinely investigated in fMRI experiments and selectively degraded by neurodegenerative diseases. The neurodegenerative selectivity for certain dynamic brain patterns, or modes of function of the complex system, and the high predictive ability of GFSS mapping for all key readouts of AD suggests a fundamental role for large-scale neurodynamic physiology in AD and related disorders. This emphasizes the urgent need to ground clinical neurology and cognitive psychology in terms of computational neuroscience in order to make meaningful advances for these incurable degenerative diseases of the mind. The GFSS framework is an important step in that endeavor.

## Data Availability

The eigenimages are available for download (https://neurovault.org/collections/AXJZMEAY/). All other data and the methods used in development of figures and tables are available in the main text or the supplementary materials.

## Acknowledgments

We would like to thank Denise Reyes and Josie Williams for project administration, Arvin Arani-Forghanian and Robert Reid for helpful discussions, and most importantly, our patients and caregivers.

## Author contributions

Conceptualization: D.J., V.L, J.G-R., H.B., M.C.M., J.G., H.J., D.K., R.P., C.J., A.F., and R.R. Data curation: D.J., M.S., J.G., and H.W. Formal analysis: D.J. Funding acquisition: R.P. and C.J. Investigation: D.J., J.G-R., H.B., M.M., B.B., D.K., and R.P. Methodology: D.J., J.G., and M.S. Project administration: D.J., D.R., and J.W. Resources: V.L, R.P., and C.J. Software: D.J. Supervision: C.J. Validation: D.J., M.S, and H.W. Visualization: D.J., M.C.M., A.F., and R.R. Writing, original draft: D.J. Writing, review and editing: all authors;

## Supplementary Materials

**Fig. S1.**
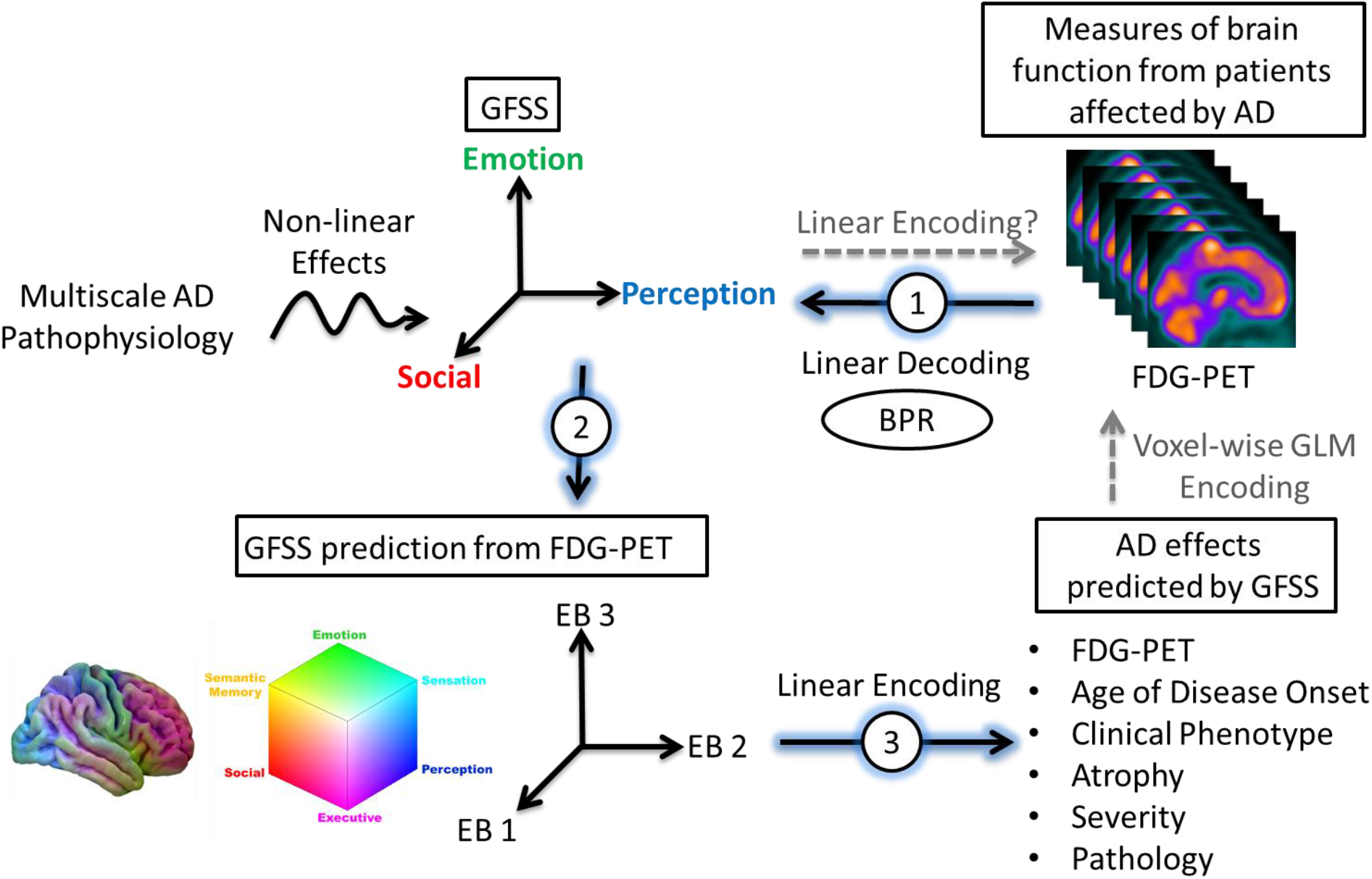
Graphical study outline. Alzheimer’s disease pathophysiology involves complex nonlinear dynamics related to the bidirectional interaction between microscale and macroscale brain organization in a manner consistent with the cascading network failure model of Alzheimer’s disease (16, 17). Unknown parameters involved in these nonlinear effects may perturb the global functional state space (GFSS) in a heterogeneous fashion leading to individual variability in the expression of this pathophysiology at the macroscale. FDG-PET is a sensitive marker of the global pattern of neurodegenerative functional disruption at the individual level. Therefore, individual variability in global FDG-PET patterns can be considered an observable parameterization of macroscale functional AD pathophysiology. Latent variable analysis of the observable variability in FDG-PET should produce a functional-anatomic mapping of the GFSS. This factor analysis assumes the underlying unseen variables are continuous and normally distributed. Therefore, the first part of this study performs a form of factor analysis computationally equivalent to the eigenface facial recognition algorithm (20), Between-subject variability Projection and Reduction (BPR), to identify spatially interpretable latent factors related to AD physiology (Fig 1). In the second part of this study, this newly identified GFSS is mapped onto functional connectivity and functional terminology (Fig 2 and Fig 3). In the third part of this study, the GFSS is used to make predictions about other manifestations of AD pathophysiology (Table 2). This allows for predictive modeling of key AD pathophysiologic manifestations in an out-of-sample multi-site study (Fig S9). This is in contrast to the more common approach of using a generalized linear model (GLM) voxel-wise, or region-of-interest based, approaches to see how these variables are encoded in FDG-PET (dashed gray upward arrow). This framework is extended to normal aging and seven dementia syndromes in the last part of the study (Fig 4).

### Participants

All participants or their designee provided written consent with approval of the Mayo Clinic Foundation and Olmsted Medical Center Institutional Review boards. All participants in the Mayo Clinic Rochester Alzheimer’s Disease Research Center and the Mayo Clinic Study of Aging that met our inclusion criteria were included in this study. As previously described (*16*), the Mayo Clinic Rochester Alzheimer’s Disease Research Center is a longitudinal cohort study that enrolls subjects from the clinical practice at Mayo Clinic in Rochester, MN. The Mayo Clinic Study of Aging is a population-based study of cognitive aging among Olmsted County, MN residents (*30*). Enrolled participants are adjudicated to be clinically normal or cognitively impaired by a consensus panel consisting of study coordinators, neuropsychologists and behavioral neurologists. Methods for defining clinically unimpaired, mild cognitive impairment and dementia in both of these studies conform to standards in the field (*31-33*). Inclusion criteria for this study consisted of 1) a Clinical Dementia Rating (CDR) global score greater than zero, 2) presence of amyloid plaques, defined as amyloid-PET standard uptake value ratio (SUVR) greater than 1.5, and 3) had high quality MRI, amyloid-PET, and FDG-PET data available for analysis. A higher more conservative SUVR cut point was used for defining amyloid-PET positivity in order to avoid contaminating the creation of this basis set with false positives (*16*). See Table 1 and Data S1 for more details on the participants included in this study.

### Structural Magnetic Resonance Imaging

As previously described (*16*), MRI was performed on one of three compatible 3T systems from the same vendor (General Electric, Waukesha, WI, USA). A 3D magnetization prepared rapid acquisition gradient echo (MPRAGE) structural imaging sequence developed for the Alzheimer’s Disease Neuroimaging Initiative (ADNI) study was acquired (*34*). All images were acquired using an 8-channel phased array head coil. Post-processing to correct for gradient distortion correction and processing has been validated in multiple studies, shown to give consistent stable results in ADNI data, and geometric fidelity after correction is independent of scanner (*35, 36*). Parameters were: TR/TE/T1, 2300/3/900 msec; flip angle 8°, 26 cm field of view (FOV); 256 × 256 in-plane matrix with a phase FOV of .94, and slice thickness of 1.2 mm. These MPRAGE parameters have been held invariant since approximately 2008. This structural MRI was used for preprocessing PET data.

### PET Acquisition and Preprocessing

The amyloid-PET imaging was performed with C-11 Pittsburgh Compound B (*37*) and FDG-PET with F-18 fluorodeoxyglucose. PET images were acquired using 1 of 2 PET/CT scanners (DRX; GE Healthcare). A computed tomography scan was obtained for attenuation correction. These images were usually acquired on the same day with 1 hour between amyloid-PET and FDG-PET acquisitions. Subjects were prepared for FDG-PET in a dimly lit room, with minimal auditory stimulation. Amyloid-PET images consisted of four 5-min dynamic frames from 40 to 60 min after injection. FDG-PET consisted of four 2-min dynamic frames acquired from 30 to 38 min after injection. PET sinograms were iteratively reconstructed into a 256 mm FOV. The pixel size was 1.0 mm and the slice thickness 3.3 mm. Standard corrections were applied.

The global amyloid-PET SUVRs were calculated as previously described (*38*). The FDG-PET image volumes of each subject were coregistered to the subject’s own T1-weighted MRI scan, using a 6 degree-of-freedom affine registration with mutual information cost function. Each MRI scan was then spatially normalized to an older adult template space (*39*) using a unified segmentation and normalization algorithm (*40*) with transforms applied to co-registered FDG-PET images. These spatially normalized images were then intensity normalized to the pons and spatially smoothed with a 6 mm full-width half-maximum Gaussian kernel.

### Between-subject variability projection and reduction

The unsupervised machine learning algorithm, Between-subject variability Projection and Reduction (BPR), was designed to capture pathophysiologic information present in between-subject variability in a disease parameter of interest. The singular value decomposition (SVD) at the heart of the data reduction portion of the algorithm is widely used and interpretable, but other methods could be used depending on the framing of the problem at hand. The goals of this algorithm also motivate data preprocessing decisions that focus on between-subject variance within the class being studied rather than variance in the observed modality under investigation or variance relative to classes not being studied. This algorithm conceptualizes multivariate medical data from an individual as representing a particular parameterization of a (patho)physiological process of interest and uses within-class individual differences in this parametrization to define a high dimensional parameter space that contains a smaller dimensional subspace manifold that describes global features of the disease generating processes of interest. This lower dimensional subspace can be isolated in many ways, but ideally the dimensionality reduction technique used would retain interpretability in order to facilitate understanding of the pathophysiology of interest and be able to meaningfully place new subjects into the learned subspace and make predictions about clinical variables of interest (Fig S1).

In the present study, we assume that macroscale glucose uptake patterns in cognitively impaired individuals with amyloid plaque deposits represent a parameterization of macroscale AD pathophysiology. We then isolated the between-subject variability of interest to this study from these preprocessed FDG-PET scans in the following way. The preprocessed FDG-PET images are three-dimensional arrays of voxel intensities that correspond to SUVR values in a standard template space. Taking only the voxel intensities that fall within the set of voxels, *V*, that have a greater than 15% probability of being gray matter in template space, this three-dimensional array can be reduced to a one-dimensional vector, ***Ψ***, with 150,468 elements at our image resolution, defined by V. To isolate subject effects, each element is non-parametrically standardized by the median, 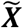 and interquartile range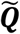 for that element across subjects 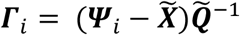 (see Fig 1 for surface renderings of 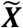 and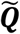). Let the set of these standardized vectors, with 150,468 elements per image, be **Γ**_1_, **Γ** _2_, **Γ** _3_ … **Γ** _M_, where M is the number of participants studied (M = 423). Subject-wise centering of each image is represented by the vector 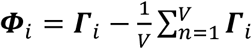. This can then be used to represent the individual differences of interest in the brain images between each image pair, or between subject variance, by calculating the subject-wise M by M matrix L,

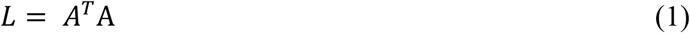

where the matrix A = [**Φ**_1_ **Φ**_2_ … **Φ**_M_]. This high-dimensional projection of individual differences can be represented as an eigendecomposition, using the singular-value decomposition *L* = *vεv*^*T*^, such that the M eigenvectors, ***v***_*i*,_ of *L*, determine the linear combination of the M set of FDG-PET images that produce image space eigenvectors, ***u***_*l*_, or eigenbrains given that they can be ordered into a three-dimensional configuration corresponding to the original brain images, as previously described for the eigenfaces facial recognition algorithm for two-dimensional facial recognition (*20*):

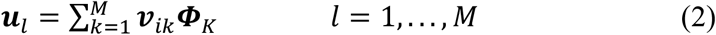

This was demonstrated while considering that the eigenvectors ***v***_*i*_ of *A*^*T*^A such that

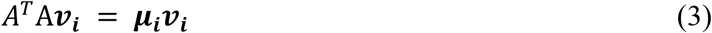

multiplying both sides by *A*,

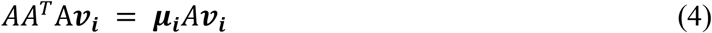

it is shown that ***Av***_*i*_ are the eigenvectors of the larger dimensional covariance matrix (150,468 by 150,468) in image space, *C* = *AA*^*T*^. This algorithm demonstrates how individual differences in macroscale multivariate patterns in brain images can be mapped back into the original image space in the form of a compact lower-dimensional basis-set of eigenbrains (EBs). This allows for a highly interpretable understanding of the parameterization of a disease processes affecting the individuals included in the analysis.

**Fig. S2.**
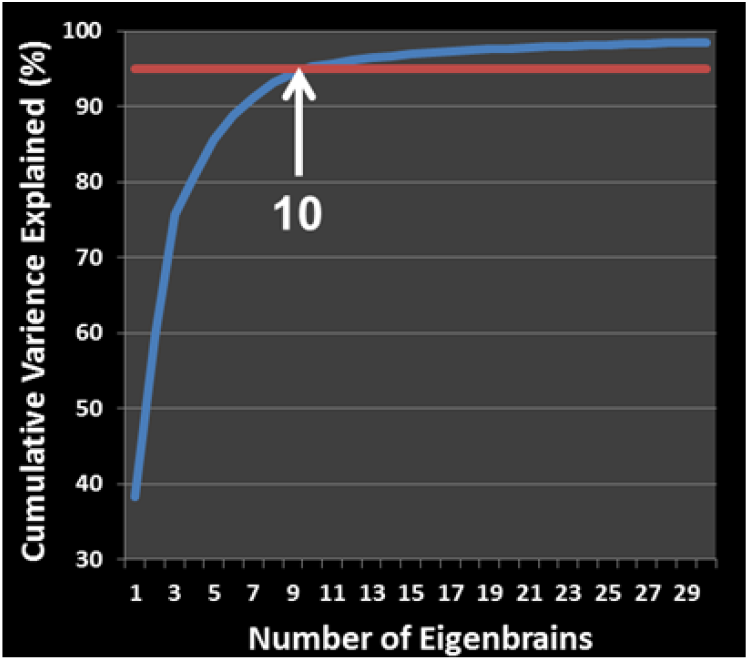
Cumulative percentage of variance explained for the first 30 eigenbrains (blue curve). The first 10 eigenbrains (arrow) explain 95% (red line) of the variance in this dataset.

The first 10 EBs (see Fig 1 for surface renderings) explained 95% of the variance in the dataset (Fig S2). Using only these 10 EBs, ***u***_*i*_, and the eigenvectors ***v***_*i*_, of *L*, as a subject-level weight, an individual FDG-PET scan can be estimated, ***Ψ***^***est***^, from a linear combination of EBs in following way:

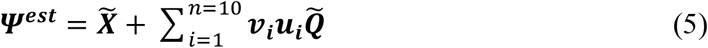

An example of an estimated image using only these 10 EBs relative to the original image is presented in Fig S3. Using additional EBs adds additional structural information, but this does not appear relevant to quantifying dysfunction in the GFSS or enhance predicative ability (see section below, *Out-of-sample predictions in ADNI*).

**Fig. S3.**
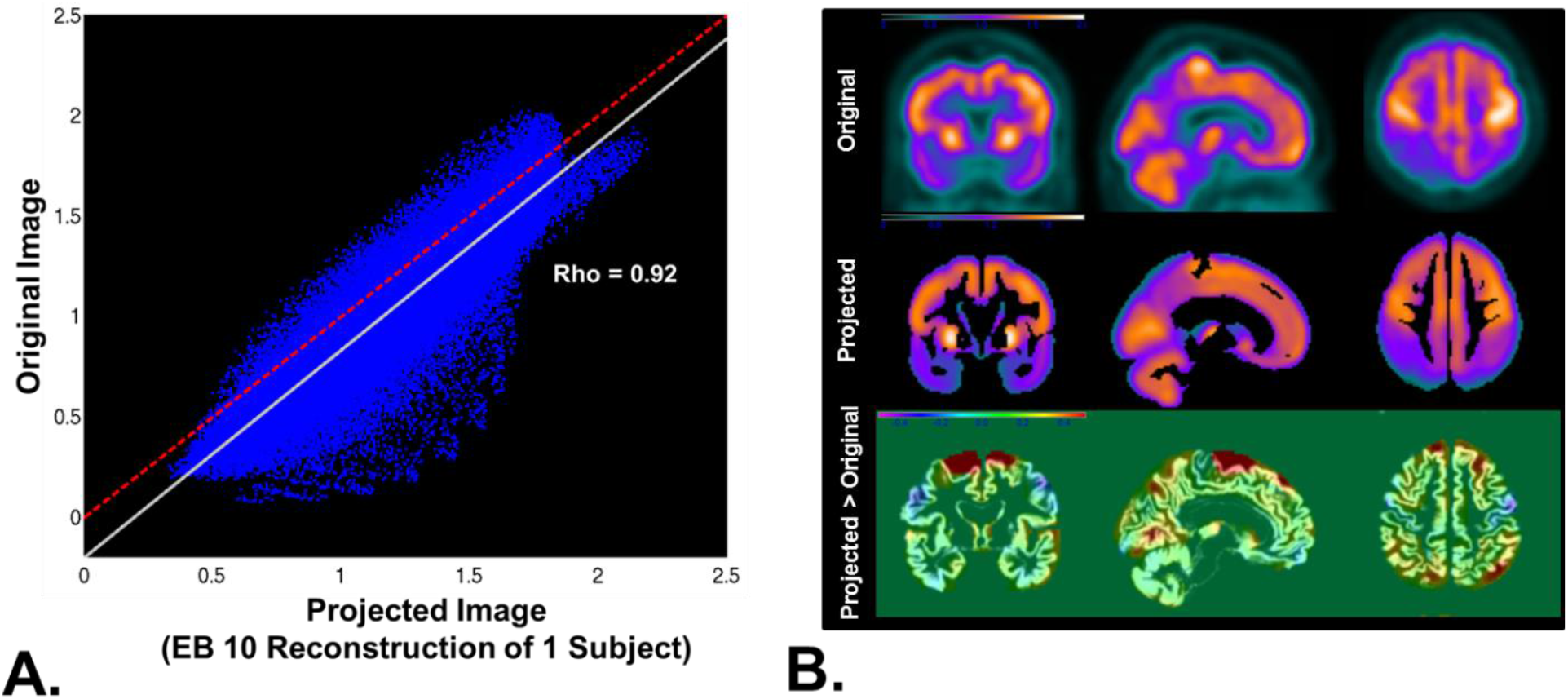
**A)** The voxel intensities in the original FDG-PET scan from one subject are plotted versus the voxel intensities of the projected images reconstructed from a 10-EB model. Red dotted line is the line of identity. The solid white line is the least square fit of the data points. **B)** Orthogonal slices of the original images (top), projected image (middle), and the difference image between the two overlaid on the gray matter segmentation (bottom). The regions in red in the difference image indicates regions that are present in the projected images but are not in the original image. These regions correspond to anatomic variations in the subject’s gray matter and are not related to global metabolic patterns.

In order to determine the robustness of this algorithm to place an unseen image into this same GFSS mapping, we iterated the algorithm 423 times leaving out each subject exactly once and estimated the subject level weights, ***v***_*i*_, for the left out subject using the first 10 EBs, ***u***_*i*_, and the associated singular values, *ε*_*i,i*_, derived from the remaining 422 subjects. These estimates were then compared to the derived values from the original run that included all 423 subjects. The set of subject-level weights, ***v***_*i*_, for an unseen image, ***Γ***_*m*_, for each of the 10 EBs, ***u***_*i*_, was calculated in the following way:

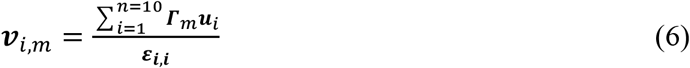

The concordance between the original values and the estimated values was assessed using the absolute value, given that the sign is indeterminate and may change on a given iteration (Fig S4). The method demonstrated a robust performance with Kendall’s coefficient of concordance approaching 1, indicating near complete agreement between the full model and the estimates obtained for the unseen left out subjects using equation *6*. To assess the sample-related bias of the basis-set produced by this data-set, we generated 500 bootstrapped samples and calculated the first 10 EBs per sample and compared the correlation of the absolute values of the EB images produced to the EBs from the original model. All 10 EBs appeared to be robust to sample variation with relatively more variation seen in the EBs explaining less than 5% of the variance in the original data (Fig S5).

**Fig. S4.**
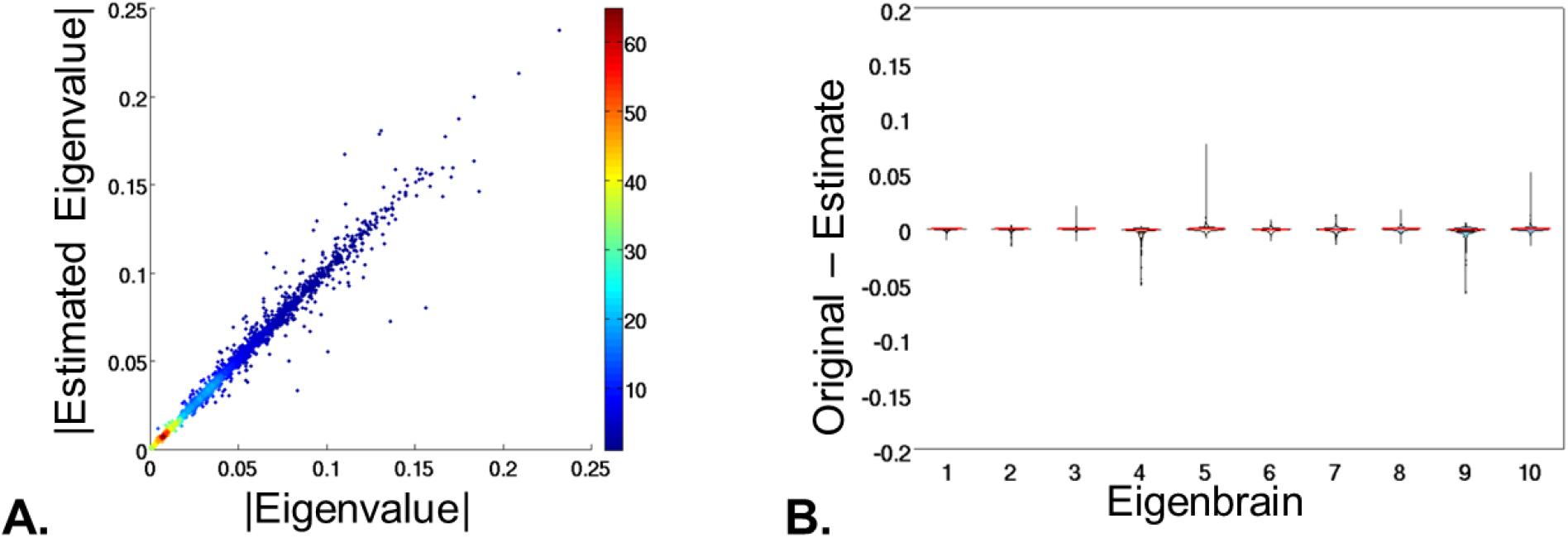
Leave-one-out cross-validation of eigenvalue estimates. **A)** The absolute values of the eigenvalues estimated from a leave-one-out model are plotted versus the absolute values of the eigenvalues obtained from the full model (point density is encoded in the colormap). This shows highly consistent results for the eigenvalues whether they were calculated in the full model or estimated after leaving that subject out (Kendal’s W approaching a value of 1). **B)** Violin plots by eigenbrain of the difference between the eigenvalues estimated in the original full model subtracted from the values estimated during the leave-one-out runs demonstrating similarly high reproducibility of eigenvalues across all eigenbrains.

**Fig. S5.**
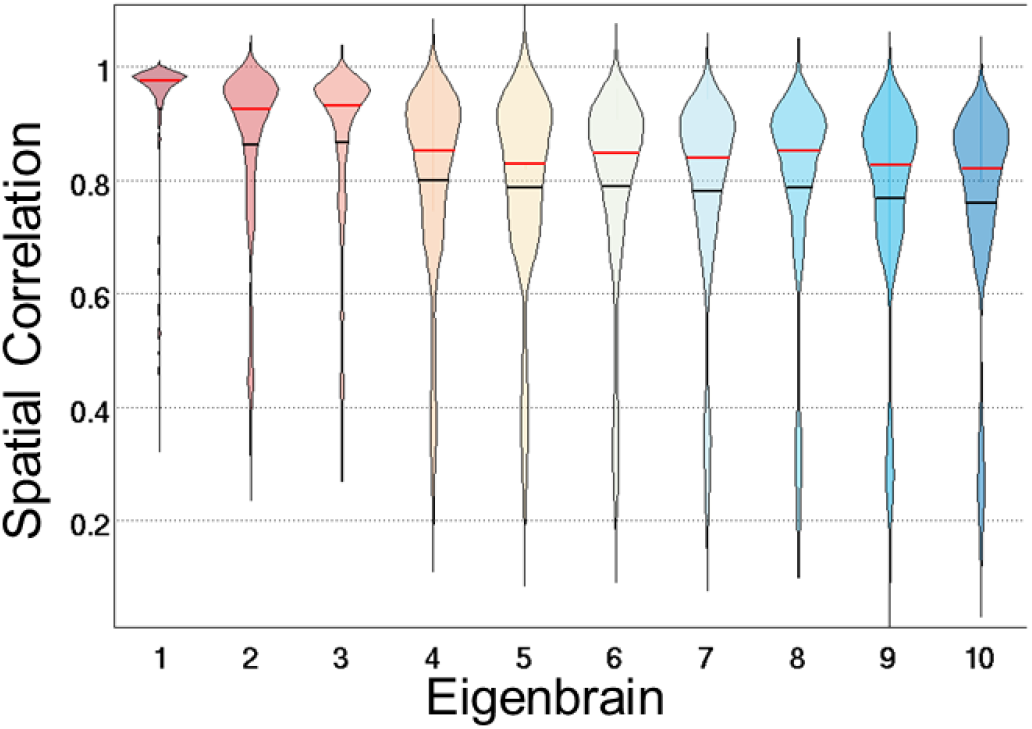
Violin plots of the spatial correlation between the eigenbrains estimated from the original full model and 500 bootstrapped samples (mean in black and median in red). All eigenbrains were robust to sample variation. The first three eigenbrains, that explained 75% of the variance in the original sample, were more stable than the remainder of the eigenbrains (EB4-10), that explaining 20% of the variance in the original sample.

We further tested the effect of sample selection on the eigenbrain maps by performing the same BPR procedure on a large dataset of FDG-PET images spanning the age and degenerative disease spectrum (N = 4,448). We found that the first three eigenbrains and eigenvalues were replicated in this larger dataset, but EB1 and EB2 switched order in terms of variance explained in the larger dataset (Fig S6). Using only a cohort of young cognitively normal participants, we were not able to replicate the eigenbrains derived from these cohorts.

**Fig. S6.**
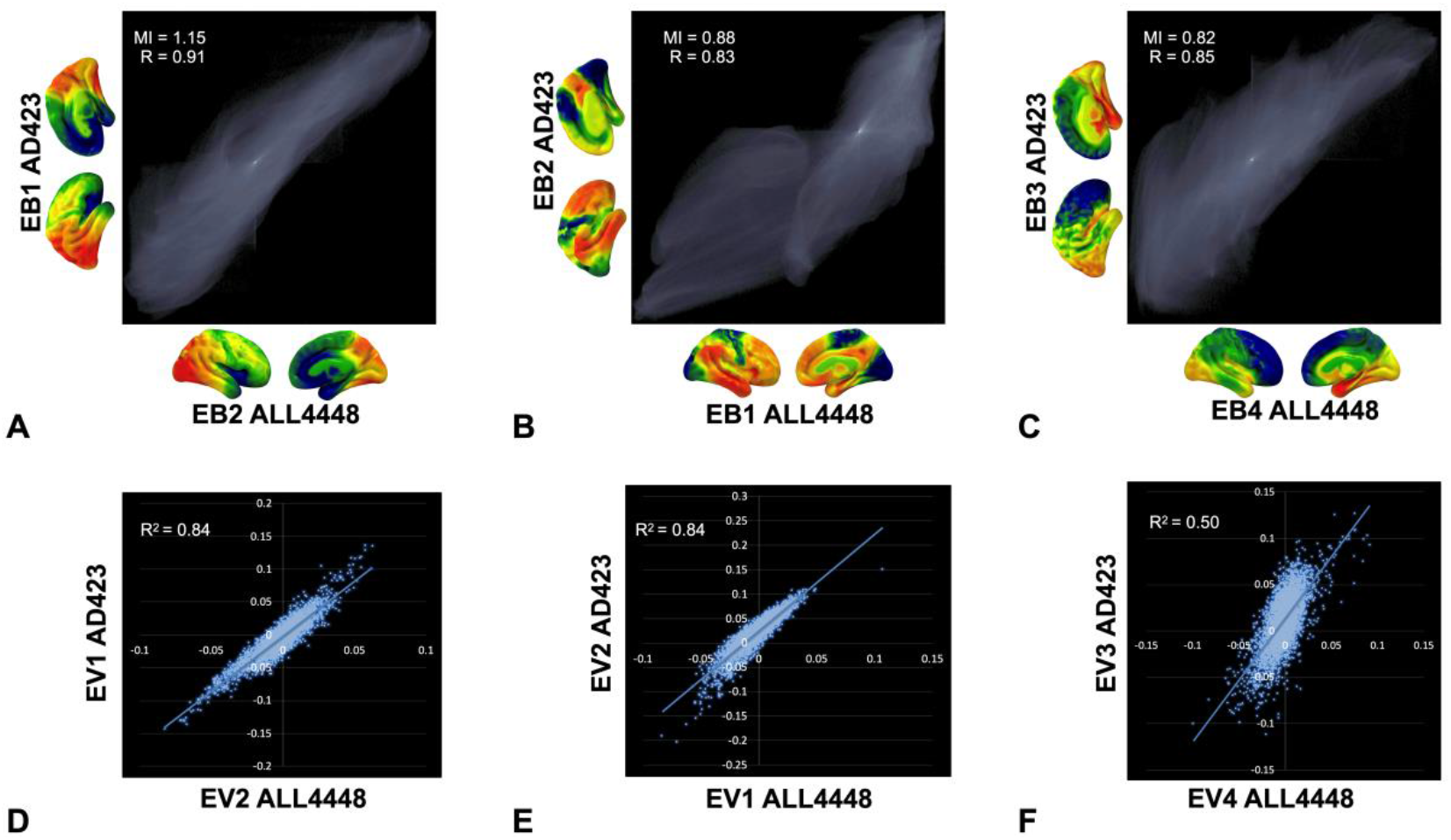
Replication of GFSS defined in the AD spectrum across a large dataset of FDG-PET images. A-C) Joint histograms between the eigenbrains defined using the AD spectrum (AD423) versus a larger FDG-PET dataset spanning the age and dementia continuum (ALL4448). The mutual information (MI) and correlation coefficient (R) is inset. D-F) Scatter plot and linear regression for the associated eigenvalues for their respective eigenbrains.

We also explored the effect of non-linear approaches to defining the manifold and found no compelling evidence not to use the linear solution provided by the PCA approach (Fig S7).

**Fig. S7.**
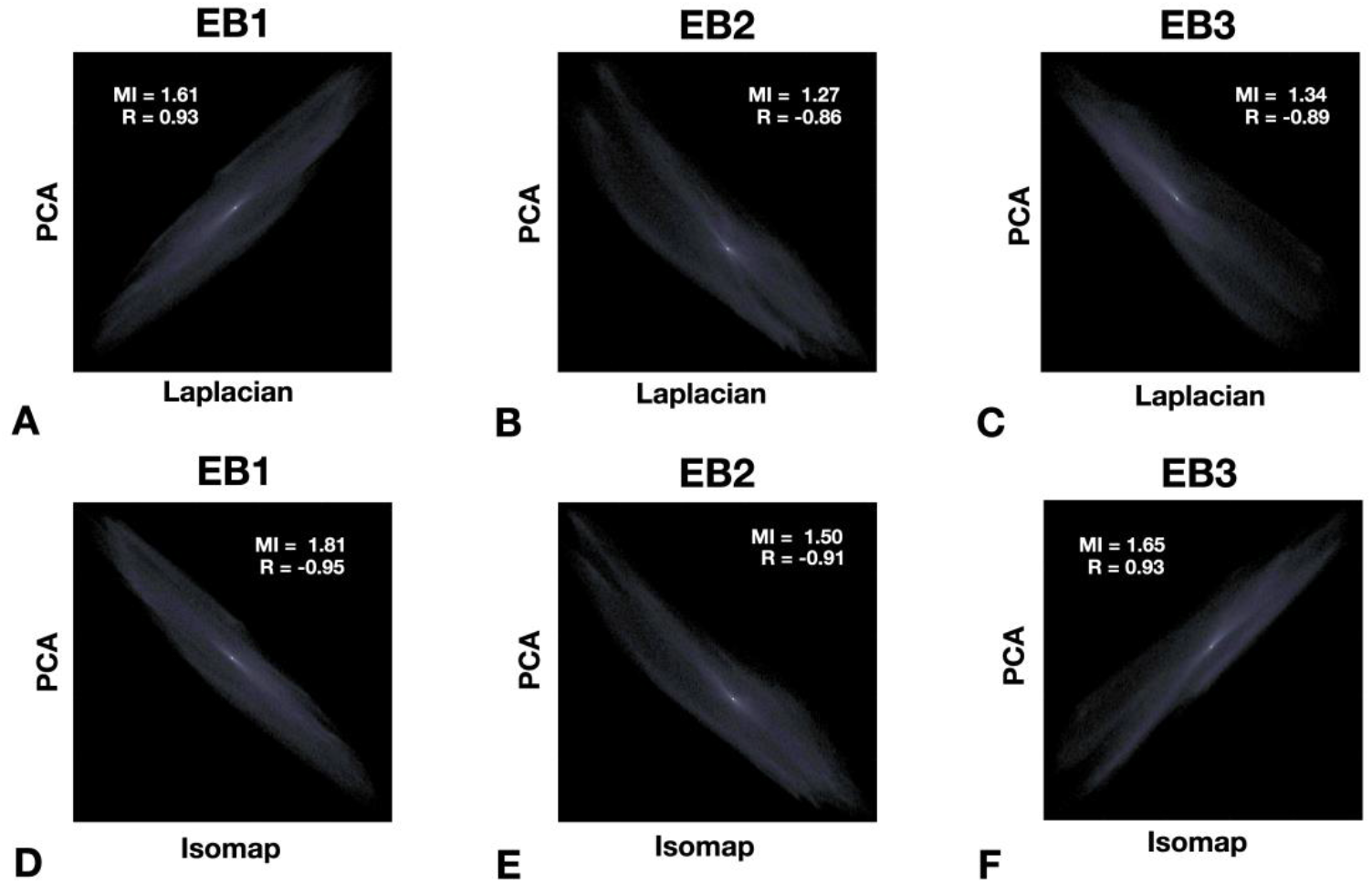
Comparing eigenbrains derived from PCA to Laplacian eigenmaps and isomaps. Joint histograps comparing the first three eigenbrains derived from PCA to Laplacian eigenmaps (A-C) and isomaps (D-F). All three methods produced similar manifolds with high mutual information (MI).

### Out-of-sample predictions in ADNI

To evaluate the out-of-sample predictive ability of the GFSS for key measures of the effects of AD pathophysiology (i.e., age, glucose uptake, cognition, and disease severity), we used the publically available multisite data from the Alzheimer’s Disease Neuroimaging Initiative (ADNI) database (adni.loni.usc.edu). This multisite database is well suited for testing the ability of the GFSS to make predictions about individual subjects with biomarker evidence of AD pathophysiology. The focus of this validation analysis was on predicting an individual’s age based on their metabolic mapping to state space. Age prediction is a strong validation of the GFSS mapping given the association between age and clinical phenotypes (*41*), pathologic phenotypes (*42*), and global tau deposition patterns (*16*). We analyzed the data from 410 ADNI subjects (Table 1) with FDG-PET scans, CDR greater than 0, and positive amyloid PET imaging as defined by previously established ADNI cut-point (1.11 for whole cerebellum referenced AV45 data) (*43*).

In this dataset, the FDG composite summary used in ADNI to summarize AD-like patterns of hypometabolism (*44*) is not associated with age (Fig S8A). However, once the eigenvalues for each of the first 10 EBs are calculated from an individual’s FDG scan, the GFSS model fits in the Mayo data (Table 2) can be used to accurately predict the age of an individual (Fig S8B) and the FDG composite score (Fig S8C). The predictive ability of the GFSS models in the ADNI data is not substantially improved with additional EBs being included in the models for FDG composite, age, disease severity, or cognition (Fig S9).

**Fig. S8.**
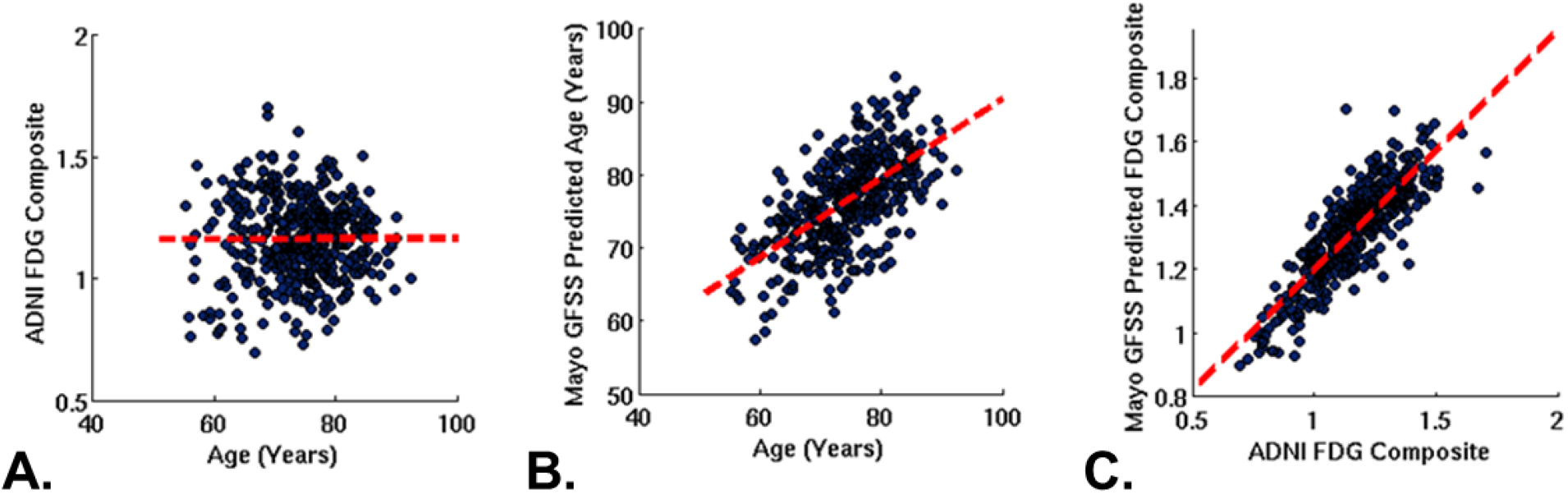
Global functional state space predictive modeling. **A)** The ADNI FDG composite score is not associated with age. **B)** Using the FDG-PET to place the subjects into the 10-dimensional GFSS allows for accurate prediction of the subjects age using models fits from the Mayo data (Table 2). **C)** The GFSS also allows for an accurate prediction of the subjects FDG-PET composite. See Fig S9 for a range of correlation values by number of eigenbrains used in the predictive model.

**Fig. S9.**
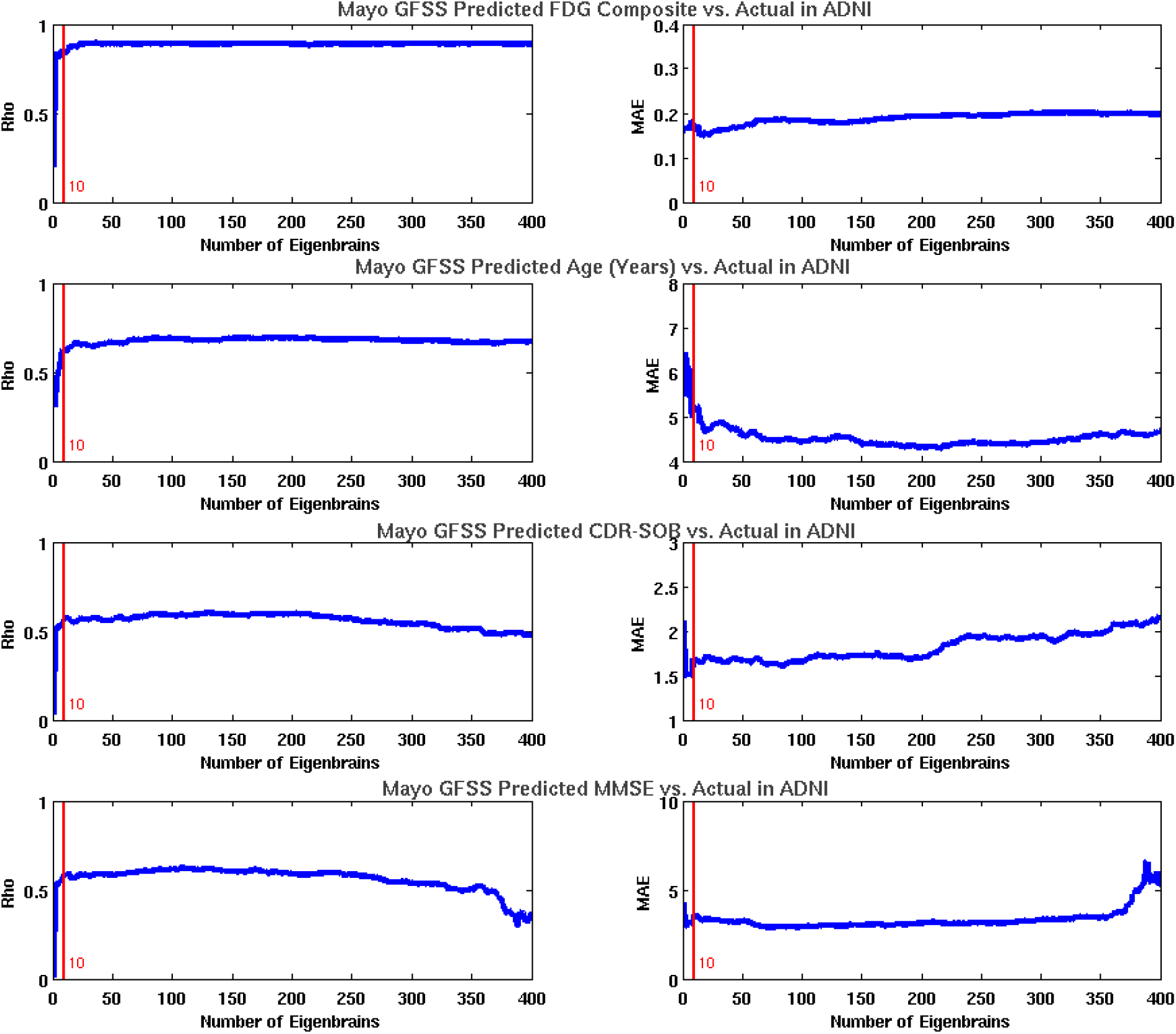
Plots of model order effect on global functional state space predictive modeling of FDG, age, severity, and cognition in ADNI. The Pearson correlation coefficients between the actual and predicted values are plotted versus the number of eigenbrains used in the predictive model for each of the variables (left). The mean absolute error (MAE) of the prediction is plotted versus the number of eigenbrains used in the predictive model for each of the variables (right). The near optimal performance of the 10-D model is highlighted by the red vertical bar in each plot.

### FDG-PET eigenbrains and large-scale functional organization of the brain

We used the NeuroSynth database (www.neurosynth.org) (*21*) and the recently described (*24*) principle gradient of macroscale functional organization (available at https://neurovault.org/images/24346/) to map our FDG-PET derived EBs to patterns of functional connectivity and functional terminology. We first calculated the voxel-wise Pearson correlation between the principle gradient of functional connectivity and EB2 and found a high correlation (r = 0.82) (Fig 3A). Next we compared a NeuroSynth topic terms (*22*) based decoding of EB2 and the principle gradient of functional connectivity. Feature terms were derived from the 50 set of topic terms (v4). Of the 50 available, 28 terms captured coherent cognitive terms spanning the theoretical range of the GFSS and mirrored the range evaluated by Margulies, et al. (2016) (*24*) and are used in further analysis. The decoding of all 28 topic terms is available in Table S1. The decoding analysis produces a Pearson correlation between the unthresholded EB and the unthresholded topic term meta-analysis images (see the FAQs section here for details: http://neurosynth.org/decode/?neurovault=308). The topic term decoding of EB2 was similar to the same analysis performed on the principle gradient of macroscale functional organization (r = 0.86) in that at one extreme were regions serving concrete primary sensory/motor functions and at the other end were abstract processes involving transmodal regions (Fig 3B). The same decoding of EB1 however revealed brain regions involved in processing external visual information were at one extreme and brain regions associated with evaluating internal mental and physical states (e.g., emotions, pain, and sustenance) were on the other extreme. EB3 was divided into brain regions involved in fluid executive control (e.g., response preparation, working memory, and response inhibition) with highly learned perceptual categories (e.g., faces, objects, and sensory perception) that can rely on feedforward control of previously learned models on the opposite extreme. The decoding weights for each of the topic terms for EB1-3 were used to associate functional terminology with the points in the three-dimensional GFSS (Fig 3C). The points in this plot were color-coded treating each EB decoding as a channel in a RGB color scheme (EB1 = Blue, inverted polarity EB2 = Red, EB3 = Green). This same RGB color-coding was done voxel-wise using the spatial loadings of EB1-3 so that a complete functional-anatomical mapping could be visualized on a brain rendering (Fig 3D). The same color-coding is then used for the eigenvalues for individual subjects included in this study (Fig 4).

## Statistical Methods

A combination of MATLAB-based (Mathworks Inc., Natick, MA, USA) and R-based (http://www.R-project.org) software packages were used to perform all statistical analysis. When comparing cohort characteristics, Kruskal-Wallis one-way ANOVA was used for continuous variables and chi-squared tests were used for categorical variables. Multiple linear regression predictive models were used to for dependent variables in Table 2, the first 10 eigenvalues were used as predictors.

---

**Table S1.** NeuroSynth Topic Term Decoding.

| Summary Term | Neurosynth Topic Term | EB1 | EB2 | EB3 | EB4 | EB5 | EB6 | EB7 | EB8 | EB9 | EB10 | FC Gradient |
| --- | --- | --- | --- | --- | --- | --- | --- | --- | --- | --- | --- | --- |
| Langue Comprehension | 36_sentences_comprehension_language | -0.0214 | -0.3514 | -0.0445 | 0.1938 | -0.1486 | -0.0308 | -0.1483 | -0.1198 | 0.041 | 0.1262 | 0.2029 |
| Social | 17_social_participants_empathy | -0.1474 | -0.2999 | -0.0536 | -0.0518 | -0.0074 | -0.1299 | -0.05 | -0.112 | -0.0355 | -0.1087 | 0.305 |
| Memory | 12_memory_retrieval_encoding | 0.0455 | -0.2117 | 0.0812 | 0.0598 | -0.1675 | -0.1574 | -0.0157 | -0.179 | 0.0558 | -0.2862 | 0.2246 |
| Language Semantics | 44_semantic_words_word | 0.0857 | -0.1808 | -0.0092 | 0.1809 | -0.2081 | 0.0089 | -0.2206 | -0.1172 | 0.037 | 0.0724 | 0.0584 |
| Negative emotion | 40_emotional_negative_emotion | -0.384 | -0.1646 | 0.0144 | -0.2162 | -0.0603 | -0.156 | -0.0529 | -0.024 | -0.065 | -0.1671 | 0.2279 |
| Visual Attention | 41_attention_attentional_visual | 0.3479 | -0.1456 | -0.0489 | 0.0034 | 0.1125 | 0.0382 | -0.0158 | -0.0992 | -0.1699 | -0.0449 | -0.0642 |
| Language Perception | 20_reading_language_words | 0.2238 | -0.1277 | 0.0085 | 0.2152 | -0.1219 | 0.0144 | -0.2229 | -0.0865 | 0.0047 | 0.1715 | -0.0525 |
| Numerical | 42_number_numerical_arithmetic | 0.2146 | -0.1217 | -0.1044 | 0.0804 | 0.081 | 0.0111 | 0.1489 | -0.0905 | -0.0137 | -0.0469 | 0.038 |
| Working Memory | 22_memory_working_wm | 0.1447 | -0.0937 | -0.1973 | 0.128 | 0.0646 | 0.0784 | 0.1039 | -0.043 | -0.0371 | -0.0494 | -0.0089 |
| Emotional Cues | 23_emotional_faces_facial | -0.278 | -0.0789 | 0.2035 | -0.2359 | -0.1116 | -0.1975 | -0.0605 | -0.0154 | -0.0119 | -0.0852 | 0.134 |
| Reward | 29_reward_decision_risk | -0.3425 | -0.0766 | -0.0477 | -0.1049 | 0.0353 | -0.1302 | -0.0697 | 0.0833 | -0.1712 | -0.2318 | 0.1689 |
| Response Preparation | 47_conflict_response_trials | 0.153 | -0.0629 | -0.2152 | 0.0371 | 0.141 | 0.2164 | 0.029 | -0.0572 | -0.1323 | 0.0009 | -0.073 |
| Hearing | 32_speech_auditory_sounds | -0.0432 | -0.0594 | 0.0339 | 0.046 | -0.0033 | 0.0633 | -0.226 | 0.1186 | 0.0935 | 0.486 | -0.1079 |
| Facial recognition | 05_face_faces_recognition | 0.2545 | -0.0474 | 0.2877 | -0.1453 | -0.0919 | -0.2326 | -0.0967 | -0.1452 | 0.1059 | -0.0902 | 0.0137 |
| Addiction | 27_alcohol_drug_smokers | -0.2676 | -0.0183 | -0.0237 | -0.1011 | 0.0498 | -0.0901 | 0.0563 | 0.0479 | -0.0791 | -0.1689 | 0.1021 |
| Objects | 01_object_objects_category | 0.3993 | 0.0141 | 0.2736 | 0.008 | -0.1466 | -0.0874 | -0.1292 | -0.1518 | 0.1027 | -0.0631 | -0.082 |
| Sustenance State | 26_food_taste_weight | -0.2974 | 0.0304 | 0.0649 | -0.1541 | -0.0321 | -0.0737 | -0.067 | 0.1392 | -0.0825 | -0.066 | 0.0273 |
| Error Learning | 25_learning_feedback_error | -0.0106 | 0.0366 | -0.1925 | 0.087 | 0.1627 | 0.1082 | 0.0176 | 0.0657 | -0.0848 | 0.022 | -0.0229 |
| Response inhibition | 08_inhibition_responses_inhibitory | -0.1458 | 0.044 | -0.1294 | -0.1367 | 0.144 | 0.1433 | 0.0082 | 0.0714 | -0.1489 | 0.0225 | -0.0436 |
| Praxis | 00_action_actions_motor | 0.3789 | 0.0767 | 0.0181 | 0.0749 | 0.0574 | 0.2589 | -0.0023 | 0.0074 | 0.0331 | 0.2129 | -0.2198 |
| Stimulus response | 19_stimulus_responses_response | 0.0005 | 0.1154 | -0.0607 | -0.0146 | 0.1098 | 0.0358 | -0.0346 | 0.0824 | -0.1107 | -0.0155 | -0.0937 |
| Motion Perception | 11_motion_visual_perception | 0.5698 | 0.1345 | 0.1701 | -0.0178 | 0.0387 | 0.0782 | -0.0528 | -0.0468 | -0.0433 | 0.0589 | -0.2713 |
| Perception | 03_visual_auditory_sensory | 0.3943 | 0.1352 | 0.1756 | 0.0177 | -0.0245 | 0.1 | -0.2513 | 0.0209 | -0.0161 | 0.1951 | -0.2703 |
| Pain | 48_pain_painful_stimulation | -0.3524 | 0.1769 | -0.0334 | -0.1115 | 0.1219 | 0.1182 | -0.0464 | 0.2013 | 0.0024 | 0.2354 | -0.1496 |
| Directed Gaze | 15_eye_spatial_gaze | 0.4143 | 0.1954 | 0.0686 | -0.0128 | 0.0895 | 0.145 | 0.0808 | -0.0899 | -0.0235 | -0.1128 | -0.2152 |
| Somatosensory | 35_stimulation_somatosensory_tms | 0.0185 | 0.2889 | -0.0412 | 0.0011 | 0.1675 | 0.3474 | 0.0604 | 0.2183 | 0.0501 | 0.3788 | -0.3453 |
| Motor | 49_motor_movement_movements | 0.0839 | 0.4018 | -0.1742 | 0.1526 | 0.1596 | 0.3841 | 0.0687 | 0.1403 | 0.0676 | 0.3036 | -0.3354 |

**Data S1. (separate file)**

Data used for figures and tables.

